# Monocyte class switch and hyperinflammation characterise severe COVID-19 in type 2 diabetes

**DOI:** 10.1101/2020.06.02.20119909

**Authors:** Fawaz Alzaid, Jean-Baptiste Julla, Marc Diedisheim, Charline Potier, Louis Potier, Gilberto Velho, Bénédicte Gaborit, Philippe Manivet, Stéphane Germain, Tiphaine Vidal-Trecan, Ronan Roussel, Jean-Pierre Riveline, Elise Dalmas, Nicolas Venteclef, Jean-François Gautier

## Abstract

**Background:** Early in the COVID-19 pandemic type 2 diabetes (T2D) was marked as a risk factor of severe disease and mortality. Inflammation is central to the aetiology of both conditions where variations in immune responses have the potential to mitigate or aggravate disease course. Identifying at risk groups based on immuno-inflammatory signatures is valuable in directing personalised care and developing potential targets for precision therapy.

**Methods:** This study characterised immunophenotypic variation associated with COVID-19 severity in type 2 diabetes. Broad-spectrum immunophenotyping quantified 15 leukocyte populations in peripheral circulation from a cohort of 45 hospitalised COVID-19 patients with and without type 2 diabetes.

**Results:** Morphological anomalies in the monocyte pool, monocytopenia specific to quiescent monocytes, and a decreased frequency of cytotoxic lymphocytes were associated with severe COVID-19 in patients with type 2 diabetes requiring intensive care. An aggravated inflammatory gene expression profile, reminiscent of the type-1 interferon pathway, underlaid the immunophenotype associated with severe disease in T2D.

**Conclusion:** Shifts in T-cell and monocyte dynamics underpin a maladaptive response to SARSCoV-2. These alterations may impact type-1 interferon signalling which is the likely source of the hyperinflammation that increases voracity of COVID-19. These findings allow the identification of type 2 diabetic patients at risk of severe disease as well as providing evidence that the type-1 interferon pathway may be an actionable therapeutic target for future studies.

**Trial registration:** NCT02671864

**Funding:** French National Agency of Research (ANR); European Foundation for the study of diabetes (EFSD); European Research Council (ERC); Francophone Society for Diabetes (SFD)

**Brief summary:** Maladapted monocyte responses including class switch, morphological anomalies and systemic hyperinflammation put patients with type 2 diabetes at higher risk of severe COVID-19

**GRAPHICAL ABSTRACT:** 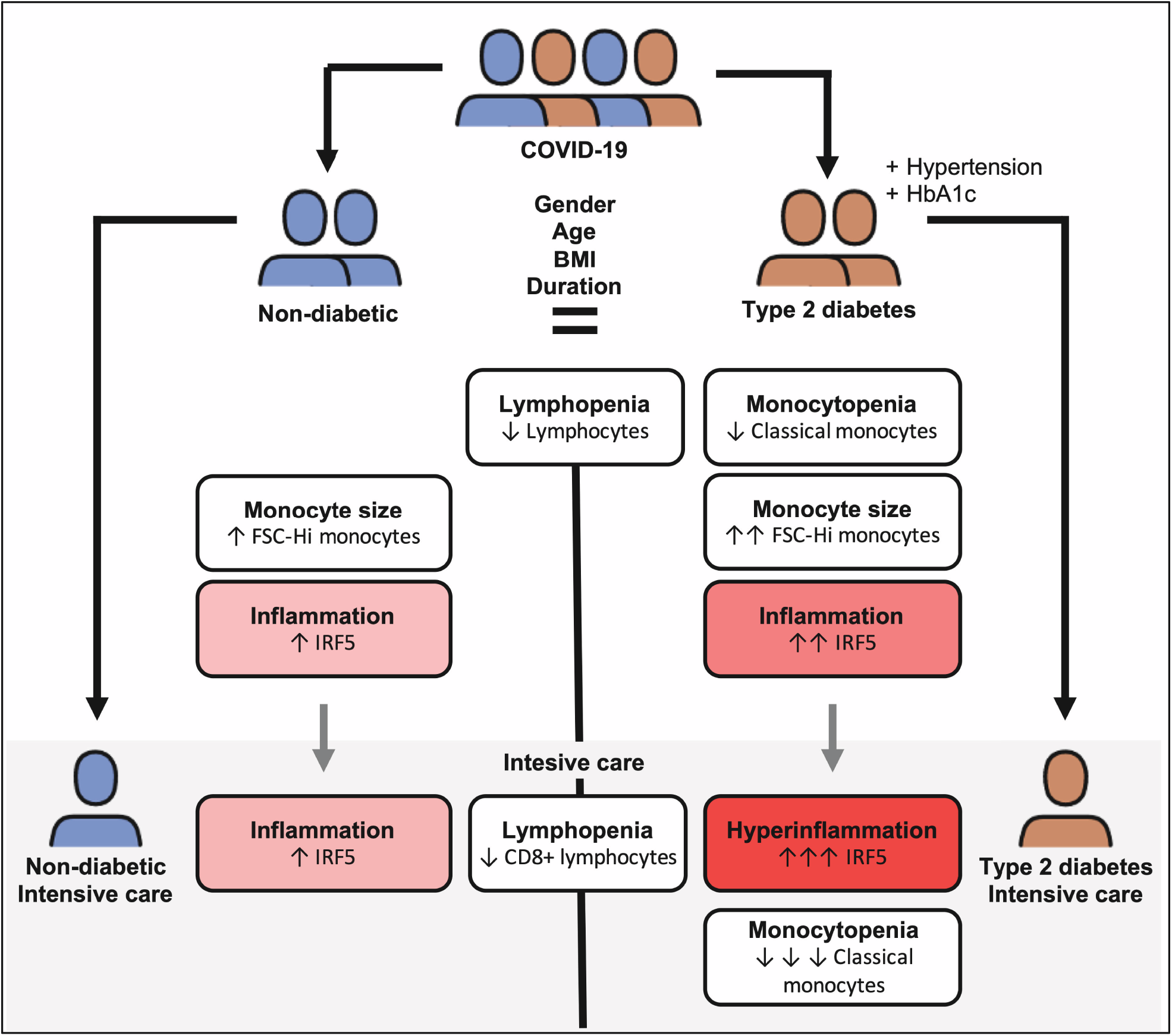

## Introduction

Coronavirus Disease-19 (COVID-19), an infectious disease caused by the severe acute respiratory syndrome coronavirus (SARS-CoV)-2 was first identified in Wuhan, China, in December 2019. SARS-CoV-2 induces strong systemic inflammation and acute injury of the lung and potentially other organs (1). As of May 18^th^ 2020, over 4.5 M cases have been reported worldwide, with an estimated mortality of 5–6% (2). Early reports stated that individuals with diabetes are at a higher risk of complications and of death from severe COVID-19 (3). Later studies indicated that factors such as age, obesity or hypertension, although commonly associated with diabetes, independently increase risk (4,5). The complex aetiologies of these conditions and of type 2 diabetes (T2D) makes deciphering the mechanisms that increase severity of COVID-19 a complex task. Potential mechanisms to explain increased severity of COVID-19 in patients with T2D, include: i) higher affinity cellular binding and efficient virus entry, ii) decreased viral clearance and iii) altered immune responses leading to hyperinflammation and aggravated “cytokine storm” syndrome. Deregulation of the immune response is a plausible hypothesis. Lymphopenia has been reported as one of the most frequent immune abnormalities in Chinese populations suffering severe COVID-19 (6). Peripheral lymphocyte counts are low, with a higher proportion of pro-inflammatory Th17 CD4+ T cells, as well as elevated cytokine levels. In addition to altered T-cell dynamics, studies revealed altered monocyte phenotypes in severe forms of COVID-19 (7). The important implication of the innate immune system in COVID-19 aetiology is also evidenced by aberrant macrophage activation at the site of infection and in peripheral tissues (8). Previous studies in T2D have highlighted abnormalities in monocyte phenotypic transition states related to disease severity and cardiovascular risk (9,10). Interactions between the heightened immuno-inflammatory states in T2D and in COVID-19 cases may well be at the root of increased susceptibility of T2D patients to severe COVID-19. Deregulation of T-cell or monocyte frequency and function may result in unchecked innate and adaptive immune responses. Such responses lead to the uncontrolled and sustained inflammation observed in severe COVID-19 and at higher risk of mortality. Herein, the immunophenotypic profile of diabetic and non-diabetic COVID-19 patients revealed specific phenotypic and morphological alteration of monocytes in T2D patients. Interestingly, the type-1 interferon sensitive transcription factor, the interferon regulatory factor (IRF)-5, is associated with altered monocyte fate in T2D. Finally, we provide evidence that a loss of CD8+ T-cells and classical monocytes concomitant to increased IRF5 expression dictate severity of COVID-19 in T2D patients.

## Results

### Type-2 diabetes is associated with decreased monocyte frequency and phenotypic alterations in COVID-19 patients

To characterise the influence of pre-existing diabetes in COVID-19 patients, we recruited a cohort of 45 COVID-19 patients from Lariboisière Hospital’s University Centre for Diabetes and its Complications including 30 patients with T2D and 15 non-diabetic patients (ND). When comparing T2D with ND patients we have no differences in clinical or demographic criteria, with the exception of increased HbA1c and hypertension in T2D relative to ND (Table 1).

**Table 1.** Characteristics of COVID-19 patients in the non-diabetic (ND) and type-2 diabetic (T2D) groups.

|  | **ND**<br>**(n=15)** | **T2D**<br>**(n=30)** | **p-value** |
| --- | --- | --- | --- |
| Age (y) | 62 (52-77) | 64 (54-77) | 0,842 |
| Sex M | 8 (53) | 23 (79) | 0,073 |
| COVID Duration at inclusion(d) | 14 (5-17) | 11 (7-17) | 0,814 |
| **Risk Factors for Severe COVID** |  |  |  |
| BMI (kg/m<sup>2</sup>) | 22 (21-22) | 28 (24-32) | 0,078 |
| Hypertension (%) | 2 (13) | 8 (62) | **0,003** |
| HbA1c (%) | 5.8 (5.4-6) | 7.8 (7.3-10.3) | **< 0.0001** |
| **Severe COVID Indicators** |  |  |  |
| Intensive Care (%) | 4 (27) | 10 (34) | 0,7384 |
| CRP (mg/l) | 138 (51-211) | 146 (86-248) | 0,725 |
| Minimum lymphocytes (10<sup>9</sup>/l) | 0.75 (0.6-1.04) | 0.9 (0.67-1.12) | 0,590 |
| D-dimers (µg/l) | 2095 (1093-3133) | 1740 (755-4505) | 0,971 |
| **Median (IQR) and Mann-Whitney for qualitative data, n (%) and chi-squared or Fisher's exact test for quantitative data. ND Non-diabetics. T2D Type 2 diabetes. HbA1c Glycated haemoglobin. CRP C-reactive protein.** |  |  |  |

To address our initial hypothesis of hyperinflammation and a dysregulated immune response to COVID-19 in patients with T2D we carried out broad-spectrum immunophenotyping of venous blood by multiparametric flow cytometry. Our approach quantifies 15 major innate and adaptive immune populations and subpopulations through lineage (CD45, CD3, HLA-DR, CD14, CD56 and CD20) and phenotypic markers (CD8, CD4, CD16, CD123, CD11c) in a single assay (Fig. S1a). Upon quantification of major immune populations, we confirmed COVID-19 associated lymphopenia (< 20 % CD45+ leukocytes) in both ND and T2D patients (Fig. 1a and Fig. S1b)(11). Unexpectedly, we found a decrease in CD14+ monocyte frequency in COVID-19 patients with T2D relative to ND patients (Fig. 1a). Of note, decreased monocyte frequency was not observed in T2D patients without COVID-19 (Fig. S1b). We did not find differences in lymphocyte, natural killer (NK), B-cell, granulocyte nor DC frequency between T2D and ND COVID-19 patients. Clinical laboratory full blood count (FBC) records corroborated these findings, with lymphopenia (< 1.5 × 10^9^/L or < 20% leukocyte count) in ND and T2D COVID-19 patients and decreased monocyte counts in T2D relative to ND COVID-19 patients (Table S1).

**Figure 1.**
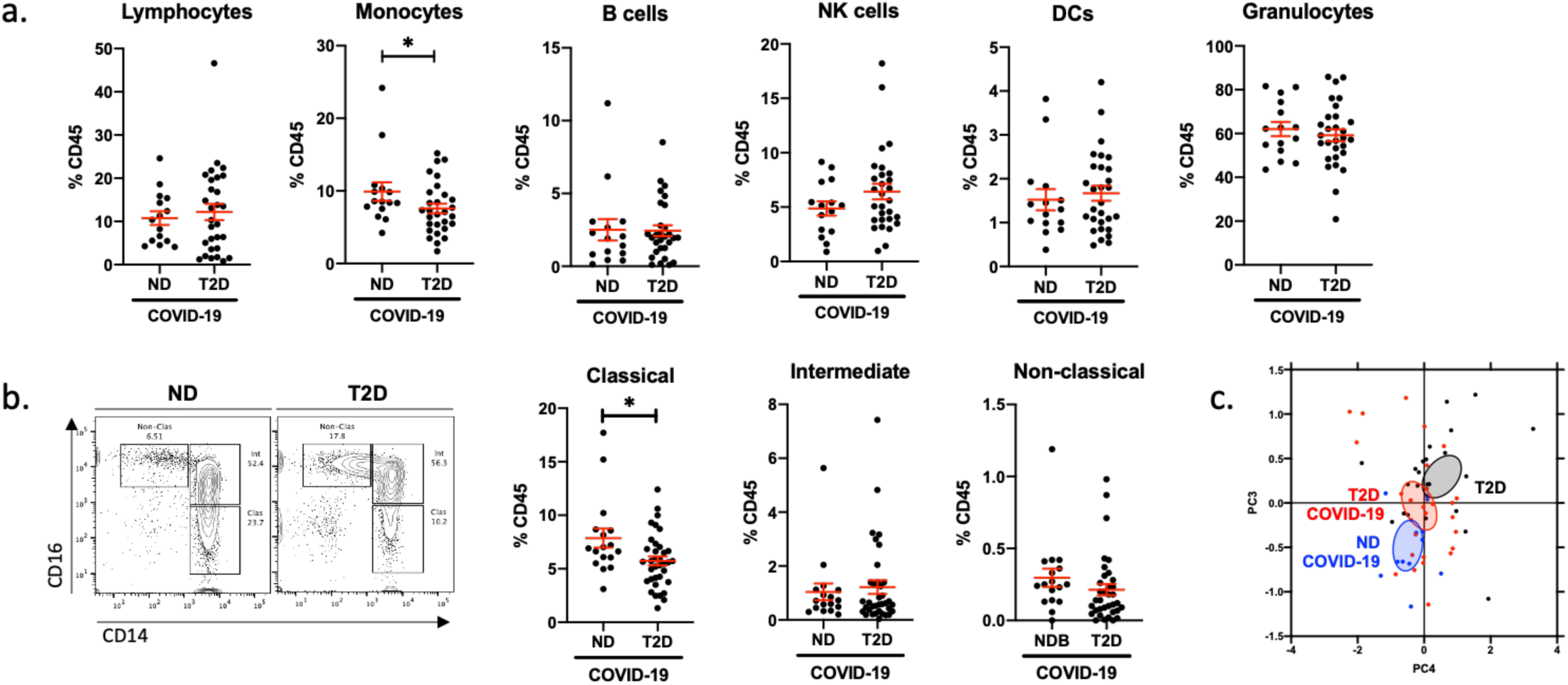
Type-2 diabetes is associated with decreased monocyte frequency and phenotypic alterations in COVID-19 patients. Flow cytometric quantificaiton of lymphocytes, monocytes, B-cells, natural killer (NK) cells, dendritic cells (DCs) and granulocytes in peripheral venous blood samples from non-diabetc (ND) and type-2 diabetic (T2D) patients with COVID-19 (a). Quantification of monocyte subpopulation phenotypes as classical (CD14^Hi^ CD16^−^), intermediate (CD14^Hi^ CD16^+^) or non-classical (CD14^Lo^ CD16^+^) (b). Principle component analysis of lymphocyte and monocyte population frequencies in ND and T2D COVID-19 patients and in T2D patients without COVID-19 (c). Data are presented as mean +/− SEM. Differences between groups were evaluated with unpaired t-test. * : p < 0.05. See also Figure S1 and Table S1.

We next quantified monocyte subtypes, to establish if a specific subtype is affected in T2D patients with COVID-19. Amongst CD14+ monocytes we found that a specific decrease in classical monocytes (CD14-Hi, CD16-) contributed to the decrease in total monocytes in T2D relative to ND COVID-19 patients (Fig. 1b). Of note, a trend (*p* = 0.07) of decreased classical monocyte frequency was also observed in T2D COVID-19 patients when compared to T2D patients without COVID-19 (Fig. S1c). We did not find any differences in the frequencies of lymphocytic, NK or DC sub-populations between ND and T2D COVID-19 patients, whilst COVID-19-associated lymphopenia was represented across all lymphocyte subtypes in ND and T2D patients (Fig. S1c).

Following these findings, we confirmed with principle component analyses (PCA) including variant immune population frequencies (lymphocyte and monocyte subpopulations), that three distinct clusters of patients emerged corresponding to ND COVID-19, T2D COVID-19 and T2D non-COVID-19 patients (Fig. 1c). When the PCA was restricted to monocytic subpopulation frequencies, ND COVID-19 and T2D COVID-19 patients also show robust separation (Fig. S1d). With multivariate ANOVA (MANOVA), integrating previously reported severe COVID-19 risk factors (gender, age, BMI, hypertension and diabetic status), we confirmed that lymphocyte and monocyte subpopulation variance is independently associated to patients with COVID-19 (p< 0.01) and T2D (p< 0.05). (Table S2).

### Morphologically altered monocytes in type-2 diabetic COVID-19 patients are associated with an aberrant inflammatory response and increased disease severity

To further characterise the monocyte pool, we quantified expression of monocyte activation markers CD16 and CD14, and of the functional pan-antigen presenting cell (APC) marker HLA-DR. We observed no difference in expression of CD16 nor HLA-DR between ND and T2D COVID-19 patients. However, T2D COVID-19 patients had decreased expression of CD14 in their monocytes (Fig. 2a and Fig. S2a). These data indicate that monocytes, between both groups of patients, have retained a similar antigen presentation capacity (HLA-DR) and have transitioned away from classical activation (CD16) at the same rate. Monocytes from patients with COVID-19 and T2D have a pronounced loss of CD14, indicating an increased rate of commitment to non-classical activation.

**Figure 2.**
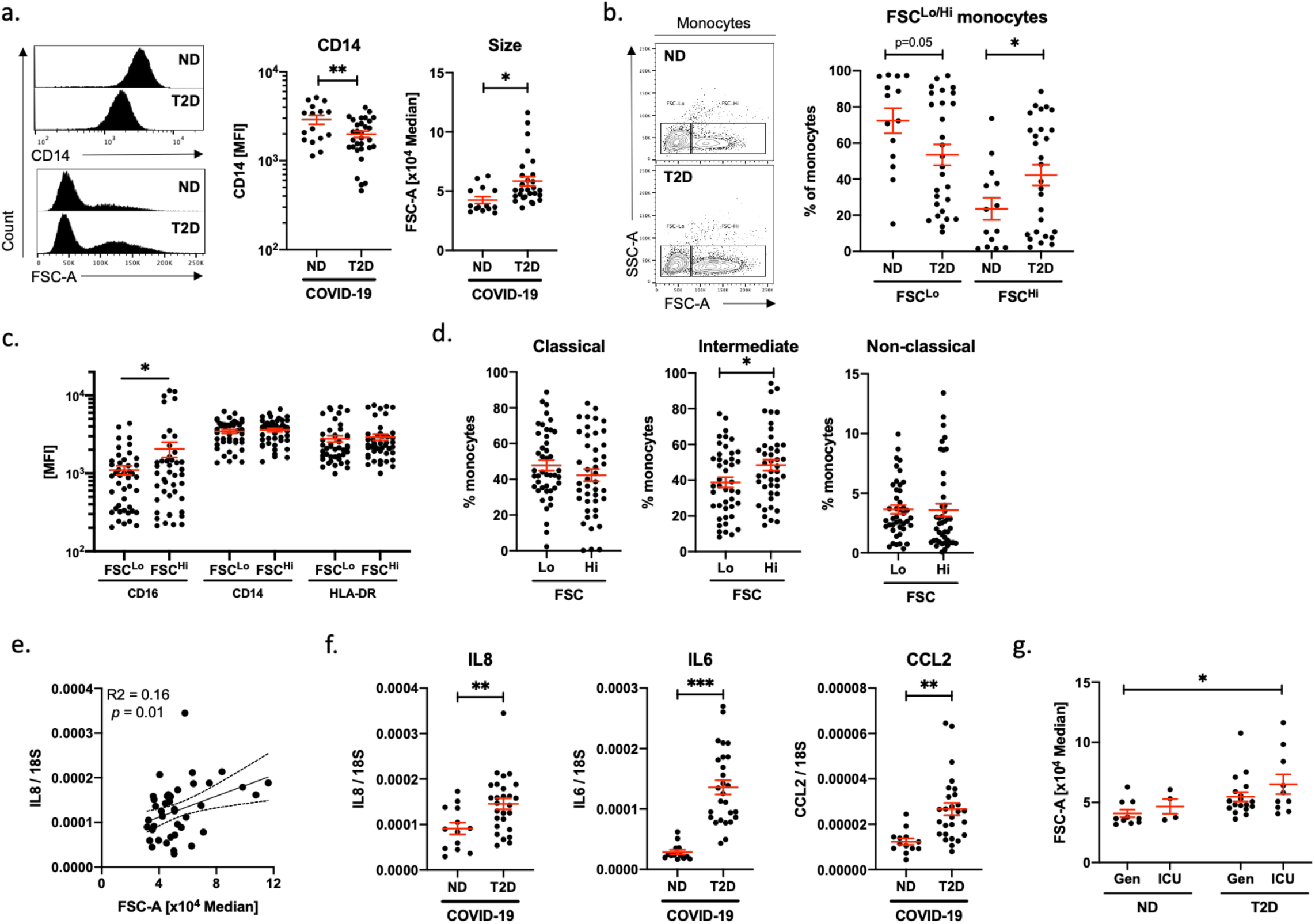
Morphologically altered monocytes in type-2 diabetic COVID-19 patients are associated with an aberrant inflammatory response and increased disease severity. Quantification of CD14 expression and size (FSC) in monocyte populations from non diabetic (ND) and type-2 diabetic (T2D) COVID-19 patients (a). Frequency of conventional (FSC^Lo^) and large (FSC^Hi^) monocytes in ND and T2D COVID-19 patients (b). Expression of CD16, CD14 and HLA-DR in FSC^Lo^and FSC^Hi^ monocytes from COVID-19 patients (c). Proportions of classical, intermediate and non-classical monocytes within FSC^Lo^and FSC^Hi^ monocytes from COVID-19 patients (d). Correlative analyses of IL8 expression in peripheral blood mononuclear cells (PBMCs) to monocyte FSC (e). IL8, IL6 and CCL2 mRNA expression in PBMCs from ND and T2D COVID-19 patients (f). Monocyte size quantified in ND and T2D COVID-19 patients treated in general wards (Gen) or in intensive care unit (ICU). Data are presented as mean +/− SEM. Differences between groups were evaluated with unpaired t-test (except for c. and d. where paired by patient). Analyses of variance (ANOVA) or covariance (ANCOVA) were used for multiple group comparisons. For correlatiive analysis Spearman’s test was carried out calculating a 2-tailed *p*-value. * : p < 0.05; ** : p < 0.01; *** : p < 0.001. See also Figure S2.

Importantly, we observed a difference in monocyte size, as indicated by forward scatter (FSC) statistics, between T2D and ND patients with COVID-19. Monocyte size was significantly higher in T2D relative to ND patients with COVID-19 (Fig. 2a). This unexpected increase in cell size was specific to CD14+ monocytes, as we next compared FSC statistics in CD14 and CD3 bifurcate gates, and found no differences between T2D and ND COVID-19 patients in CD14-, CD3+ nor CD3- cells (Fig. S2b). We next quantified the frequencies of the two distinct populations of FSCLo (conventional) and FSC-Hi (large) monocytes in our cohort and corroborated a higher frequency of FSC-Hi monocytes in T2D relative to ND COVID-19 patients (Fig. 2b and Fig. S2c).

To characterise this atypical population of FSC-Hi monocytes, we quantified expression of other known lineage markers (CD20, CD56, CD3) independently of patient status. We found no differences in the expression of non-monocyte markers (CD20, CD56, CD3) between FSC-Lo and FSCHi monocytes (Fig. S2d). Monocyte activation and functional marker analyses revealed increased expression of CD16 in FSC-Hi monocytes relative to FSC-Lo monocytes (Fig. 2c). CD16 positivity is associated with intermediate and non-classical monocyte class switch. We next quantified the proportions of each monocyte class within FSC-Hi and –Lo populations and found higher proportions of CD16+ intermediate monocytes in the FSC-Hi population relative to FSC-Lo (Fig. 2d).

Increased phenotypically switched FSC-Hi monocytes and decreased CD14 expression in the T2D monocyte pool (Fig. 1b) indicate that patients with COVID-19 and T2D present a hyper-inflammatory phenotype due to exacerbated loss of classical monocytes. To evaluate the extent of the inflammatory response associated with monocyte size and T2D in COVID-19 we carried out gene expression analyses on PBMCs from the patients included in our cohort. Monocyte size was significantly and positively correlated to expression of the pro-inflammatory marker interleukin (IL)-8, while other inflammatory markers (IL6, CCL2) trended towards positive correlation to monocyte size (Fig. 2e and Fig. S2e). When COVID-19 patients are stratified based on diabetic status, the expression of multiple inflammatory markers is drastically increased in presence of T2D (Fig. 2f) Of note, ND patients with COVID-19 have a higher level of expression of inflammatory markers relative to non-COVID-19 patients with T2D (Fig. S2f).

Given the above findings, we hypothesised that monocyte size would be of prognostic value at presentation. Thus, we stratified patients based on admission to intensive care unit (ICU) or care in general wards for non-critical patients (Gen). We found monocyte size to be increased in T2D patients admitted to the ICU relative to ND patients that did not require ICU admission (Fig. 2g). When comparing ICU to Gen, disregarding diabetic status, we did not detect a statistically significant difference in monocyte size (Fig. S2g).

### IRF5 expression is associated with monocyte activation and morphological adaptation in COVID-19 patients

Given the described aetiology of COVID-19 and the patterns of inflammatory marker expression, we hypothesised that the type-1 Interferon response is engaged, and likely dysregulated in T2D patients. The transcriptional mediator of this pathway is the Interferon Regulatory Factor (IRF)-5, controlling expression of such genes as IL6 and IFNB1. It is highly expressed in the myeloid compartment, is canonically responsive to viral stimuli (12,13), and is involved in regulating metabolic inflammation (14,15).

We quantified mRNA expression of IRF5 and of its target type-1 interferon, IFNB1, in PBMCs from our cohort. We found IRF5 and IFNB1 to be induced in ND COVID-19 and with an exaggerated increase in T2D COVID-19 patients (Fig. 3a and Fig. S3a). Furthermore, expression of IFNB1 correlates to that of IRF5, indicating likely dependent expression in this sample (Fig. 3a). Then, we quantified IRF5 expression in circulating cells by FACS and found its highest levels to be in monocytes and other populations with myeloid compartments (DC, NK) (Fig. 3b and Fig. S3b). Whilst IRF5 expression was not variant in the total monocyte pool between patient groups (Fig. 3c and Fig. S3c); we found its expression to increase throughout monocyte activation (Fig. 3d). IRF5 expression is increased in intermediate monocytes of T2D patients relative to ND patients, with a similar trend in non-classical monocytes (Fig. 3e and Fig. S3d).

**Figure 3.**
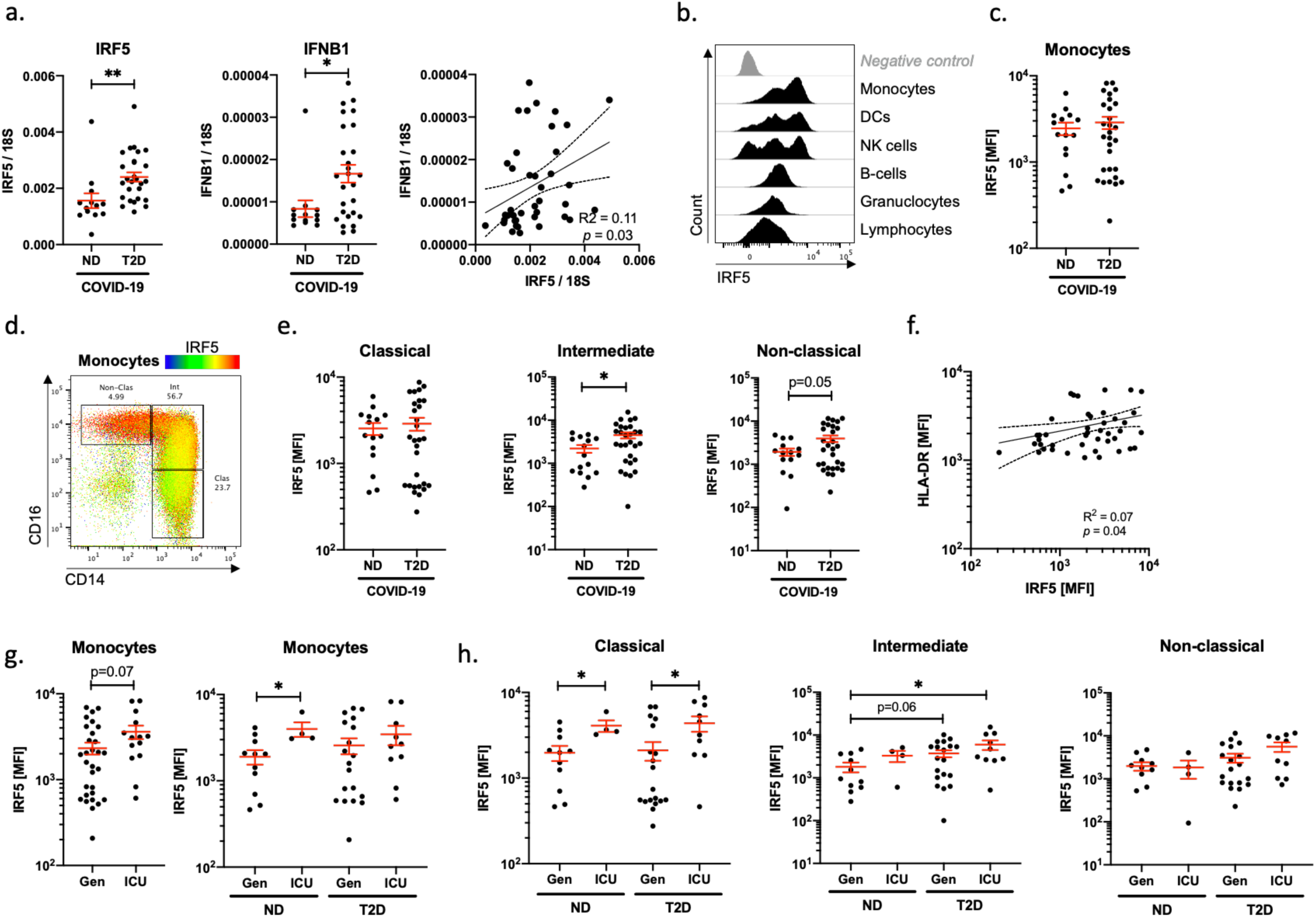
IRF5 expression is associated with monocyte activation and morphological adaptation in COVID-19 patients. Quantification of IRF5 and IFNB1 mRNA expression in peripheral blood mononuclear cells of non-diabetic (ND) and type 2 diabetic (T2D) COVID-19 patients (a). Histograms of IRF5 expression on different populations analysed by fow cytometry on venous blood samples (b). IRF5 median fluorescence intensity (MFI) in monocytes of ND and T2D patients with COVID-19 (c). IRF5 expression overlaid onto monocyte phenotypic gating (d). IRF5 expression (MFI) in monocyte subtypes from ND and T2D COVID-19 patients (e). HLA-DR and IRF5 expression in monocytes from COVID-19 patients (f). IRF5 expression in monocytes of ND or T2D COVID-19 patients admitted to intensive care unit (ICU) or treated exclusively in general wards (Gen) (g). IRF5 expression in monocytes of ND or T2D COVID-19 patients admitted ICU or treated exclusively in Gen (f). Data are presented as mean +/− SEM. Differences between groups were evaluated with unpaired t-test. Analyses of variance (ANOVA) was used for multiple group comparisons. For correlative analysis Spearman’s test was carried out calculating a 2-tailed *p*value. * : p < 0.05; ** : p < 0.01. See also Figure S3.

Correlative analyses to monocyte phenotypic and functional markers revealed a positive correlation between IRF5 and HLA-DR, with no correlation to CD14, CD16 nor FSC (Fig. 3f and Fig. S3e). These data indicate that IRF5 does not directly regulate monocyte class switch nor morphological changes, however a dependent relationship exists between IRF5 and HLA-DR. IRF5 may therefore impact antigen presentation capacity or other functions associated to HLA-DR.

To evaluate whether the inflammatory response mediated by IRF5 could infer protection or severity of COVID-19, we stratified our patients based on their admission to ICU. In the monocyte pool we observed increased expression in patients admitted to ICU, significant only amongst ND COVID-19 patients (Fig. 3g). Interestingly, in monocyte subpopulations we observed a specific signature associated with ICU admission taking into account patient diabetic status. Classical monocyte expression of IRF5 is specifically increased in patients admitted to the ICU, whereas intermediate monocyte expression of IRF5 is only increased in T2D patients admitted to ICU (Fig. 3h).

### Severity of COVID-19 is associated with loss of CD8+ lymphocytes and loss of classical monocytes in patients with T2D

Despite the majority of ICU admissions from our cohort being patients with T2D; the proportions of ICU admissions within ND patients was comparable (26.7% of ND vs and 34.5% of T2D; Fig. 4a). To decipher the immunophenotypic and inflammatory signatures more specifically associated with COVID-19 severity in T2D, we compared mild-moderate and severe-critical cases of COVID-19 in this patient group.

**Figure 4.**
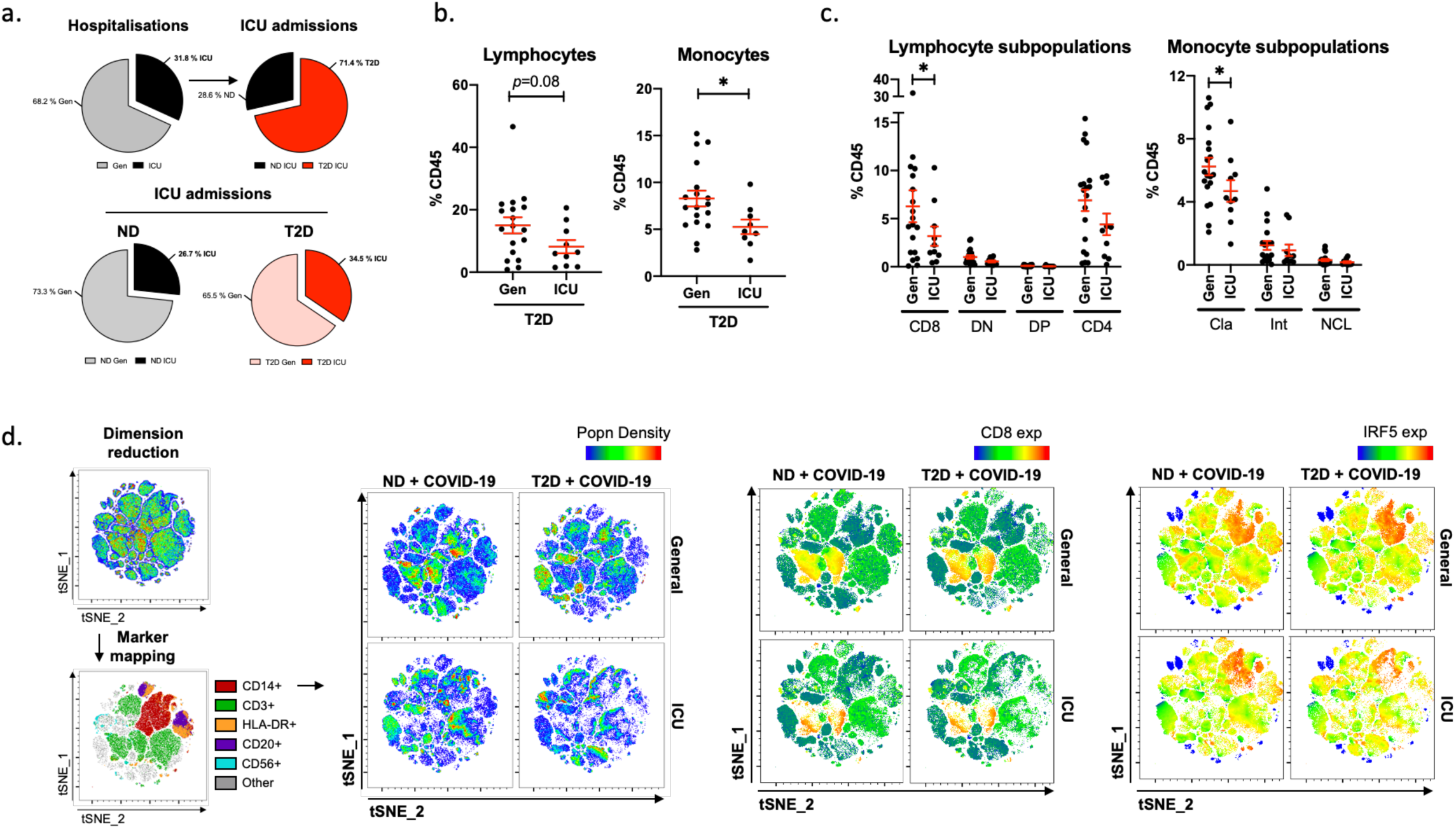
Severe COVID-19 is associated with loss of CD8+ lymphocytes and loss of classical monocytes in patients with T2D. Pie charts of COVID-19 hospitalisation that were exclusively treated in general wards (Gen) or that required intensive care (ICU). Proportions of patients admitted to ICU that were non-diabetic (ND) or with type-2 diabetes (T2D) (a). Proportions of lymphocytes and monocytes in peripheral venous blood from T2D patients with COVID-19 either treated in Gen or requiring ICU admission (b). Phenotypic analysis of lymphocyte and monocyte subpopulations in peripheral venous blood from T2D COVID-19 patients treated in Gen or requiring ICU treatment (c). tSNE mapping of all cytometric acquired data with projections of population density, CD8 or IRF5 IRF5 expression. Maps are of representative profiles from each group (d). Data are presented as mean +/− SEM. Differences between groups were evaluated with unpaired t-test or non-parametric Mann-Whitney U test. * : p < 0.05. See also Figure S4.

When patients with T2D are stratified based on ICU admission, we observe a significant decrease in monocyte frequency and a trend to decreased lymphocyte frequency in T2D patients requiring ICU admission relative to those treated in general wards (Gen) (Fig. 4b). The decrease in lymphocyte and monocyte proportions is corroborated by hospital issued FBCs (Fig. S4a). Quantification of immune subpopulations revealed concomitant decreases in CD8+ T-cells and classical monocytes in ICU, the latter of which confirms monocytopenia, specific to classical monocytes, as a hallmark of COVID-19 infection in patients with T2D (Fig. 4c). A further loss of classical monocytes is indicative of severity in T2D. No other populations in circulation were significantly affected in the ICU group (Fig. S4b and Fig. S4c).

We next applied unsupervised analyses to our cytometry data. Consisting of dimensional reduction by the tSNE algorithm (16), marker mapping by projecting known lineage markers to the tSNE map and evaluating variation between patients. Visual analysis of tSNE maps confirmed the monocyte and lymphocyte dynamics associated with T2D and with COVID-19 severity; notably a marked loss of CD14+ monocytes and CD8+ lymphocytes in T2D patients with severe COVID-19; versus a specific loss of CD8+ lymphocytes in ND patients with severe COVID-19 (Fig 4d). When projecting IRF5 expression over the tSNE maps, we confirm monocyte-specific expression, maintained at a high level despite marked monocytopenia in severe COVID-19 with T2D.

### Discussion

2020 has been marred by the emergence of SARS-CoV-2 affecting over 4.5 M people globally. The resulting Coronavirus Disease (COVID)-19 has placed unprecedented strain on health services and claimed over 300 K lives (2). With diabetic patients accounting for approximately 10% of COVID-19 deaths urgent mobilization of diabetologists and diabetes researchers is required to define criteria that confer risk. An example of such valuable work has found that appropriate glycaemic control is a key determinant of survival amongst T2D patients with COVID-19 (3). Our current study investigates inflammation and immune cell dynamics as possible mechanisms conferring risk of severe COVID-19 in patients with T2D.

In COVID-19 patients with or without T2D, we carried our broad-spectrum immunophenotyping of 15 leukocyte populations and subpopulations in peripheral circulation, evaluating changes associated with diabetic status and disease severity. We found that T2D patients with COVID-19 are characterized by monocytopenia, specific to quiescent monocytes. Monocyte loss was accompanied by morphological alterations and a hyperinflammatory expression profile consistent with the type-1 interferon response. These phenomena, alongside a marked loss of CD8+ T-cells in peripheral circulation characterise severe COVID-19 in pre-existing T2D.

### Lymphopenia, monocytopenia and phenotypic variation in COVID-19 with pre-existing T2D

In keeping with recent reports, we confirmed lymphopenia as a hallmark of COVID-19 (6). What we were astounded to observe was the added monocytopenia in patients with T2D. To our knowledge, ours is the first report of monocytopenia specific to T2D patients suffering a coronavirus infection, and such is likely to be a particularity of COVID-19. Monocytopenia has been previously associated with acute infection (17). Whilst in our current study, duration of COVID-19 was not different between groups and thus monocytopenia is likely specifically related to patient diabetic status. Of note, monocyte loss detected by flow cytometry is corroborated by clinical FBC and thus could serve as an additional COVID-19 screening tool for diabetology practice, with lymphopenia as an established indicator of severity in the general population.

Beyond the quantitative analyses, we identified an exaggerated phenotypic class switch of monocyte subpopulations in COVID-19 patients with T2D. As monocytes undergo activation they transition from their classical phenotype to intermediate and non-classical phenotypes gaining expression of CD16 and gradually lowering expression of CD14 (18). We demonstrate an increased rate of transition from the classical phenotype, towards CD16+ intermediate monocytes, and an accelerated loss of CD14 towards the non-classical phenotype in T2D. Both chronic inflammation and viral infection (in particular influenza viruses) are associated with switching monocyte class (19,20). Importantly, monocyte expression of CD16 increases potency of inducing CD4+ T-cell polarisation compared to quiescent (classical) monocytes that are CD16- (21). This could in-part explain the hyperinflammatory response in patients with T2D that have accelerated monocyte activation and a marked loss of quiescent monocytes.

A specific loss of classical monocytes may indeed be due to increased apoptosis when confronted with a viral challenge, as has been reported in *in-vitro* modelling (19). Alternatively, monocytes in T2D patients may be primed to transition towards their activated damage-seeking state more rapidly than in ND patients. Of note the accumulation of intermediate and non-classical monocytes is associated with increased rate of infection and a largely maladaptive response (19,22,23). Moreover, non-classical monocytes have recently been reported to important drivers of pulmonary hypertension in T2D patients (24). Taken together these data are coherent in explaining, in part, increased risk of severe disease or mortality in COVID-19 patients with T2D.

### Morphological alterations and hyperinflammation in COVID-19 patients with T2D

A recent pre-print reported that monocyte morphological alterations are associated with COVID-19 (7). These alterations consisted of increased monocyte size (FSC^Hi^ monocytes) in a subset of COVID-19 patients. Interestingly, altered monocyte size has also been reported in infections of influenza virus (19). A major novelty of our investigation is the characterisation of these morphologically altered monocytes in T2D. We corroborate the recent pre-print report of FSC^Hi^ monocytes in COVID-19, and we demonstrate an increased frequency of FSC^Hi^ monocytes in T2D. Moreover, we demonstrate that monocyte size is associated with a hyperinflammatory gene expression profile in PBMCs and with admission to ICU of patients with T2D. Surprisingly little is known with regards to the regulation of monocyte morphology, further basic research is required to decipher the natural history and functional implications of such phenomena.

### Exacerbated IRF5-linked hyperinflammation in T2D patients with COVID-19

With prior knowledge of pathways engaged in both viral and diabetic contexts we targeted a factor that controls transcription of type-1 interferons, IRF5. Previous studies have well-characterised the pro-inflammatory function of IRF5 in tissue macrophages, the progeny of circulating monocytes (12,15). In the current study we demonstrate that IRF5 expression in circulating cells is induced in COVID-19 and is increased in activated monocytes of patients with T2D relative to ND patients. Indeed, expression is correlated to that of inflammatory markers. A specific increase in activated subtypes of monocytes is in agreement with previous reports that IFR5 drives monocyte damage-seeking behaviour and differentiation to inflammatory macrophages (25,26). Moreover, increased expression of IRF5 in different compartments of the monocyte pool also marked those patients that required intensive care.

These data are in agreement with recent reports on IRF5 promoting cytokine production in response to viral challenge, notably increasing systemic inflammation without impacting viral replication (13). A further pre-print has also implicated the impairment of type-1 interferon signalling in severity of COVID-19 cases (27). Taken together, our data and previous reports indicate that basal levels of IRF5, preceding-SARS-CoV-2 infection, are dysregulated in T2D patients. Monocytopenia and rapid class switch of monocytes in T2D with COVID-19, may be the result of an exuberant viral response from an immune system primed on an inflammatory background. IRF5-linked hyperinflammation will induce eager damage-seeking behaviour, antigen presentation and cytokine release, without affecting viral replication. Thus, contributing to the cytokine storm syndrome that characterises severe COVID-19 (28).

### CD8+ T-cell loss and monocytopenia are hallmarks of severe COVID-19 in T2D

In our cohort, as in others, not all T2D patients require intensive care, approximately 10% more T2D patients required admission to ICU than ND patients. The immunophenotype of patients with T2D that required ICU admission was marked by an exacerbated monocytopenia and lymphopenia; in the case of lymphocyte phenotypes we were surprised to find that cytopenia was specific to CD8+ T-cells. A recent correspondence also highlighted a loss of CD8+ cytotoxic lymphocytes (CTLs) in patients with COVID-19, both severe and mild forms (29). The CTL population is known to play a crucial role in recognising and killing virally infected cells (30). This study by Koutsakos *et al* demonstrates that CTLs are important in conferring cross-reactivity across a number of viral strains, in effect a key target population that holds important clues for neutralising vaccine development. It is thus not surprising that those patients with under-represented CD8+ T-cells develop a more severe form of disease. A higher frequency of CD8+ T-cells and their efficient interactions with APCs, such as monocytes, may be a key process in mitigating severe disease.

Finally, unsupervised clustering of immune populations based on expression data, also mapped a marked loss CD8+ T-cells to ICU admission of both T2D and ND patients with COVID-19, as well as a loss of monocytes in T2D patients with severe disease. Importantly, when IRF5 is projected onto these clusters, we observe a sustained high level of expression specific to monocytes, even amongst populations that decrease in frequency. These data indicate that in T2D, exuberant inflammation, likely mediated by IRF5 in monocytes, maybe responsible for increased mobilisation of these cells. Their increased mobilisation decreases their abundance in peripheral circulation or at sites of interaction with lymphocytic populations. A mismatched ratio of antigen-presenting monocytes to CD8+ T-cells is a tempting mechanism for the descent from mild-moderate COVID-19 to severe-critical COVID-19. Once patients are identified as at-risk at admission, well-timed and –targeted immunosuppressive intervention may prevent the uncontrolled and persistent inflammatory response in severe or mortal COVID-19.

### Limitations

A main limitation of our study is the sample size and diversity, inclusion of veritable mild cases that did not require hospitalisation would help support and extend our conclusions with regards to the factors that increase COVID-19 severity. Increasing sample size and diversity within the T2D group would also allow the further stratification of patients based on severity of diabetes, in terms of glycaemic control or the existence of complications or comorbidities. Although the current study hypothesised that T2D itself was a risk factor through increasing systemic inflammation, and thus we aimed and achieved the recruitment of groups comparable in terms of age, BMI, gender and disease duration (as well as clinical factors that indicate severity such as CRP, D-dimers). Lastly, the “cytokine storm” syndrome was not documented in this cohort; however, several publications have very concretely established its presence in COVID-19 cases. Given the time-sensitive nature of this public health problem we privileged novel analyses to directly address the hypothesis.

## Data Availability

Information with regards to data availability on request. Prior to peer-review raw data cannot be made public. We conform to standard requirements.

## Acknowledgements

F.A. was supported by grants from the French National Agency of Research (ANR) for ANR MitoFLAME (ANR-19-CE14-0005) and the European foundation for the study of diabetes (EFSD)/Lilly grant (Characterisation of monocyte metabolism and bioenergetic responses in type-2 diabetes and risk of cardiovascular disease). N.V. was supported by grants from the French National Agency of Research (GLUTADIAB and ANGIOSAFE), the European Foundation for Diabetes (EFSD), and the European Union H2020 framework (ERC-EpiFAT 725790). Human study was performed at the Clinical Investigation Centre (Groupe Hospitalier Saint-Louis/Lariboisière, Paris) and were supported by Assistance Publique des Hôpitaux de Paris (ANR-DGOS Project AngioSafe T2D; J.-F.G., principal investigator) and ASSERADT (a non-profit patient association). E.D was supported by grants from the French and European Foundation for Diabetes (SFD and EFSD). We would like to thank Hélène Fohrer-Ting of the Centre for Cytometry, Histology and Cellular Imaging (CHIC) core facility of the Cordeliers Research Centre (CRC), for her guidance, patience, and support. The authors thank the Unité de Recherche Clinique of Lariboisière/Fernand Widal (Pr Eric Vicaut & Dr Véronique Jouis), the Délégation à la Recherche Clinique de Paris – Ile de France and the Direction de la Recherche Santé de Marseille for their administrative support. The authors also thank Nassima Haddadi, Djamila Bellili, Hanane Mersel for their technical support, nurses and patients who accepted to be involved in these studies.

## Author Contribution

FA, JBJ, MD, ED, NV and JFG designed research studies, analysed data, and wrote the manuscript. Patient inclusion was carried out by JBJ, JPR, JFG and BG. FA, JBJ, CP and NV conducted experiments, and acquired and analysed data. MD and GV carried out data and statistical analyses. BG, SG, LP, RR, JPR helped with experimental design and scientific discussion.

## Materials and Methods

### Human Populations

All consecutive patients hospitalised in April 2019 for COVID-19 infection in the dedicated University Centre of Diabetes and its Complications, Lariboisière hospital, Paris, France, were prospectively included in the study. Clinical and anthropometric data are summarised in Table 1. COVID-19 diagnosis was based on positive oropharyngeal swab RT-PCR. Exclusion criteria were type-1 diabetes, immunotherapy for previous transplantation, and treatment of COVID-19 by corticotherapy or immunotherapy before inclusion. For the analysis, patients were divided into two groups: with or without T2D. Patients with T2D are patients already known to have it before hospitalisation or patients for which diabetes has been discovered during hospitalisation (HbA1c > 6. 5% or two fasted glycemia above 7 mmol/l). The study was conducted in accordance with the Helsinki Declaration and was registered in a public trial registry (http://Clinicaltrials.gov; NCT02671864). This study was approved by local institutions and ethical committees, the Ethics Committee of CPP Ile-de-France granted approval for all individuals (Ile de France V number 15070). All patients provided informed consent indicating that they understood the nature of their participation in the study (NCT02671864). The principal investigator of this clinical trial is Prof. Gautier Jean-François.

### Data collection

Sampling, clinical data (age, gender, BMI, Diabetes Duration, history of HTA, COVID duration since 1s symptoms) and biological data (HbA1c, CRP, lymphopenia, D-dimer) were collected at inclusion in a standardised manner; events like introduction of COVID-19 specific treatments, transfer in Intensive Care Unit (ICU) and death were collected at the end of the hospitalization. Minimum lymphocyte count and maximal CRP were collected at the end of the hospitalization.

### Immunophenotyping by Flow cytometry

Blood cells were obtained from 1 mL of venous blood, after red blood cell lysis and resuspended into FACS buffer as previously described (31). After 10 min incubation with an Fc blocker (120–000–422; Miltenyi), cells were stained for surface markers with the appropriate antibodies and a Live/Dead viability dye (L34957; Thermo Fisher Scientific) according to manufacturers’ protocol. The following antibodies were used: anti-HLA-DR (AC122) and anti-CD8 (BW135/80) from Miltenyi; anti-CD14 (MΦP9), anti-CD3 (UCHT1) and anti-CD123 (7G3) from BD Biosciences; anti-CD16 (3G8), anti-CD56 (HCD56), anti-CD20 (2H7) and anti-CD11c (3.9) from Biolegend; anti-CD4 (S3.5) and anti-CD45 (HI20) from Thermo Fisher Scientific. After washing, cells were fixed and stained using the Foxp3-staining kit (00–5523–00; Thermo Fisher Scientific) according to the manufacturer’s protocol and using the anti-IRF5 (ab21689; Abcam) antibody for 1h at 4°C in the dark, followed by the donkey-anti-Rabbit-PE (12–4739–81; Thermo Fisher Scientific) secondary antibody for 20 minutes at 4°C in the dark. Acquisition was performed on a LSR-Fortessa flow cytometer (BD Biosciences) and analysed with FlowJo software (Tree Star).

### RT–qPCR analysis

RNA was extracted from blood cells (PBMCs) using the RNeasy RNA Mini Kit (Qiagen). Complementary DNAs were synthesised using super script kit (Promega). RT–qPCR was performed using the QuantStudio 3 Real-Time PCR Systems (ThermoFisher Scientific). 18S was used for normalization to quantify relative mRNA expression levels. Relative changes in mRNA expression were calculated using the comparative cycle method (2−ΔΔCt). Primer sequences were designed using Primer3 (32)(33) (http://bioinfo.ut.ee/primer3-0.4.0/) used: IL8 (F: AGACAGCAGAGCACACAAGC; and R: ATGGTTCCTTCCGGTGGT); CCL2 (F: TTCTGTGCCTGCTGCTCAT; and R: GGGGCATTGATTGCATCT); IRF5 (F: GATGGGGACAACACCATCTT; and R: GGCTTTTGTTAAGGGCACAG); IL6 (F: GCCCAGCTATGAACTCCTTCT; and R: GAAGGCAGCAGGCAACAC); IFNB1 (F: GGAAAGAGGAGAGTGACAGAAAA; and R: TTGGATGCTCTGGTCATCTTTA) and 18S (F: TTCGAACGTCTGCCCTATCAA; and R: ATGGTAGGCACGGCGACTA).

### Statistical analyses

Data are expressed as mean or median ± SD or SEM as indicated in the text, tables and figures. For quantitative data, differences between groups were evaluated with non-parametric Mann-Whitney U test, and categorical and binary variables were tested by the two-tailed Pearson chi-square test or Fisher exact test if more than 20% of the cells in the frequency tables had an expected frequency below 5. Multivariate analyses of variance (ANOVA) or covariance (ANCOVA) were used for group comparisons. Repeated measures analyses of variance (MANOVA) were employed to compare the variance of monocyte populations taking into consideration gender, BMI, hypertension and diabetic status. Statistical analyses were carried out using JMP (SAS Institute Inc, Cary, NC), XLSTAT 2014 (Addinsoft, Brooklyn, NY), Graphpad Prism (Graphpad), SPSS Statistics (SPSS corporation) and R Software 3.6.0 (http://www.r-project.org). Statistical significance was set at p< 0.05. Principal component analysis (PCA) was performed from total lymphocyte and monocytes sub-populations FACS quantification with FactoMineR R package (doi 10.18637/jss.v025.i01), and factoextra package (factoextra.bib) was used to construct graphics. Contribution of each individual was analyzed to identify outliers biasing the PCA, leading to exclusion of 4 individuals for a technical bias or hospitalisation in the ICU prior to COVID-19 infection.

## Supplementary material

**Table S1.** Full blood count of COVID-19 patients in the non-diabetic (ND) and type-2 diabetic (T2D) groups at admission to hospital.

|  | ND<br>(n=15) | T2D<br>(n=30) | P-value<br>(Mann-Withney) |
| --- | --- | --- | --- |
| Leucocytes ($10^9/L$) | 6.3 (5.5-7.8) | 6 (4.8-7.45) | 0.255 |
| Neutrophils ($10^9/L$) | 3.93 (3.23-5.27) | 3.81 (2.92-5.08) | 0.497 |
| Lymphocytes ($10^9/L$) | 1.18 (1.07-1.75) | 1.3 (0.91-1.65) | 0.860 |
| Monocytes ($10^9/L$) | 0.63 (0.48-0.76) | 0.44 (0.33-0.62) | 0.028 |

**Table S2.**
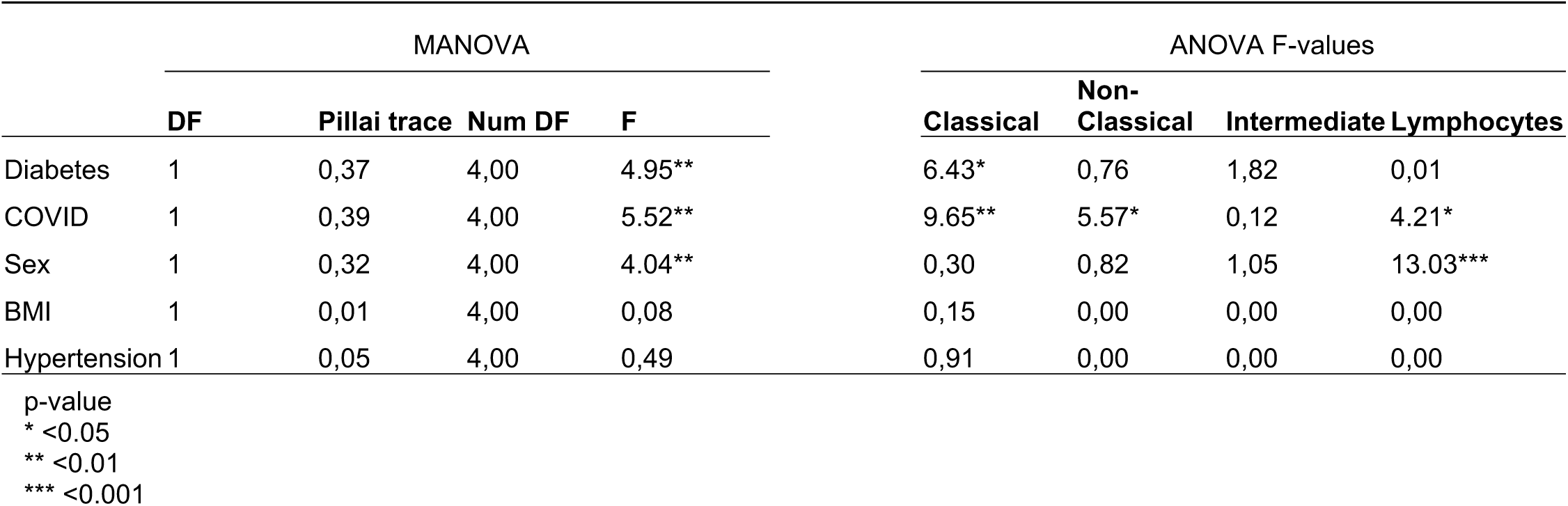
Multivariate ANOVA (MANOVA) and univariate ANOVA F-values to test differences among cell-type percent according clinical characteristics.

|  | MANOVA |  |  |  | ANOVA F-values |  |  |  |
| --- | --- | --- | --- | --- | --- | --- | --- | --- |
|  | DF | Pillai trace | Num DF | F | Classical | Non-Classical | Intermediate | Lymphocytes |
| Diabetes | 1 | 0,37 | 4,00 | 4.95** | 6.43* | 0,76 | 1,82 | 0,01 |
| COVID | 1 | 0,39 | 4,00 | 5.52** | 9.65** | 5.57* | 0,12 | 4.21* |
| Sex | 1 | 0,32 | 4,00 | 4.04** | 0,30 | 0,82 | 1,05 | 13.03*** |
| BMI | 1 | 0,01 | 4,00 | 0,08 | 0,15 | 0,00 | 0,00 | 0,00 |
| Hypertension | 1 | 0,05 | 4,00 | 0,49 | 0,91 | 0,00 | 0,00 | 0,00 |

p-value
\* <0.05
\*\* <0.01
\*\*\* <0.001

**Figure S1.**
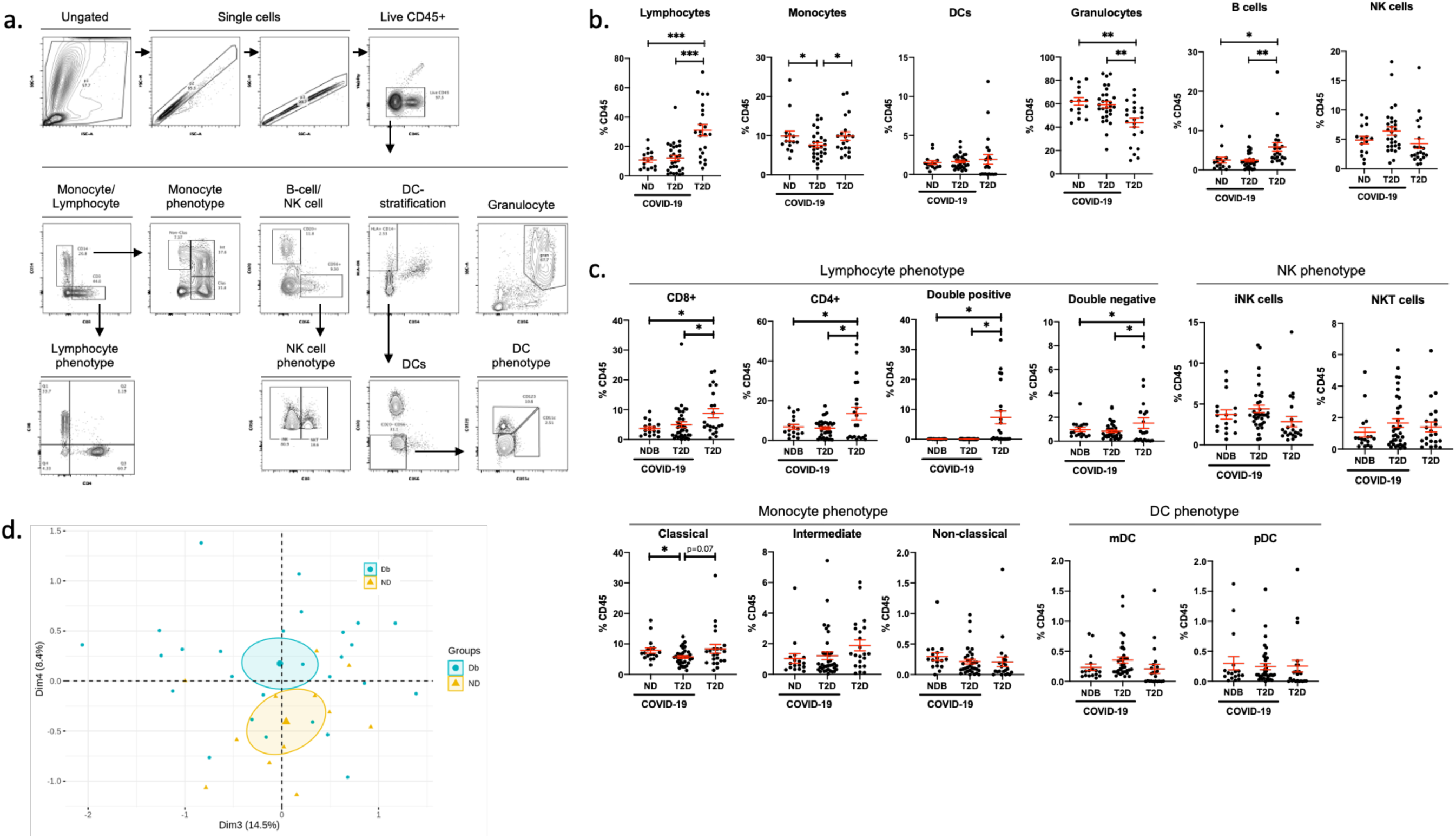
Immunophenotyping approach and subpopulations quantified in peripheral venous blood. Related to figure 1. Markers and gating strategy applied for immunophenotypic analysis of immune populations in peripheral circulation (a). Major immune populations quantified in venous blood samples from type-2 diabetic (T2D) patients and in COVID-19 patients with T2D or in non-diabetic (ND) patients (b). Subpopulations quantified in peripheral circulation of T2D patients and of COVID-19 patients with T2D or ND (c). Principle component analysis on monocyte subpopulations from ND or T2D COVID-19 patients (d). Data are presented as mean +/− SEM. Differences between groups were evaluated with unpaired t-test. *p*-value. * : p < 0.05; ** : p < 0.01; *** : p < 0.001.

**Figure S2.**
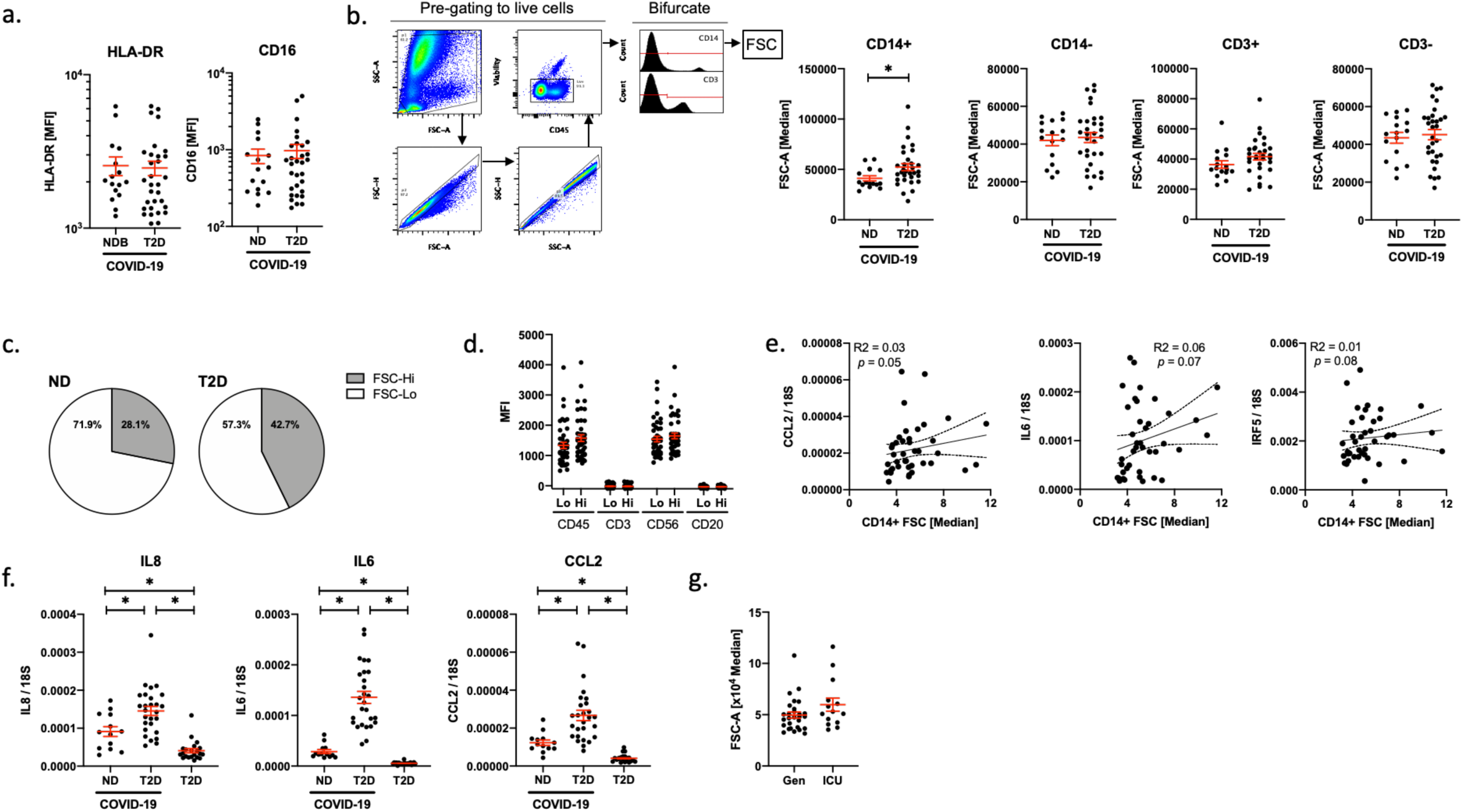
Characterisation of monocyte morphology and inflammatory cells from peripheral circulation ND and T2D COVID-19 patients. Related to figure 2. HLA-DR and CD16 expression in monocytes from ND and T2D COVID-19 patients (a). Alternative bifurcate gating strategy to evaluate monocyte and lymphocyte size (FSC) in CD14 and CD3 +/− cells from ND and T2D COVID-19 patients (b). Pie charts representing proportions of conventional (FSC^Lo^) and large (FSC^Hi^) monocytes in ND and T2D COVID-19 patients (c). Lineage marker expression in FSC^Lo^ and FSC^Hi^ monocytes from ND and T2D COVID-19 patients (d). Correlation of inflammatory marker gene expression from peripheral blood mononuclear cells (PBMCs) to monocyte size (FSC) in ND and T2D COVID-19 patients (e). Gene expression of inflammatory markers in PBMCs from ND and T2D COVID-19 patients and from T2D patients without COVID-19 (f). Monocyte size (FSC) in ND and T2D COVID-19 patients treated in general wards (Gen) or admitted to the intensive care unit (ICU). Data are presented as mean +/− SEM. Differences between groups were evaluated with unpaired ttest (except for d. where pairing was by patient). Analyses of variance (ANOVA) or covariance (ANCOVA) were used for multiple group comparisons. For correlative analysis Spearman’s test was carried out calculating a 2-tailed *p*-value. * : p < 0.05

**Figure S3.**
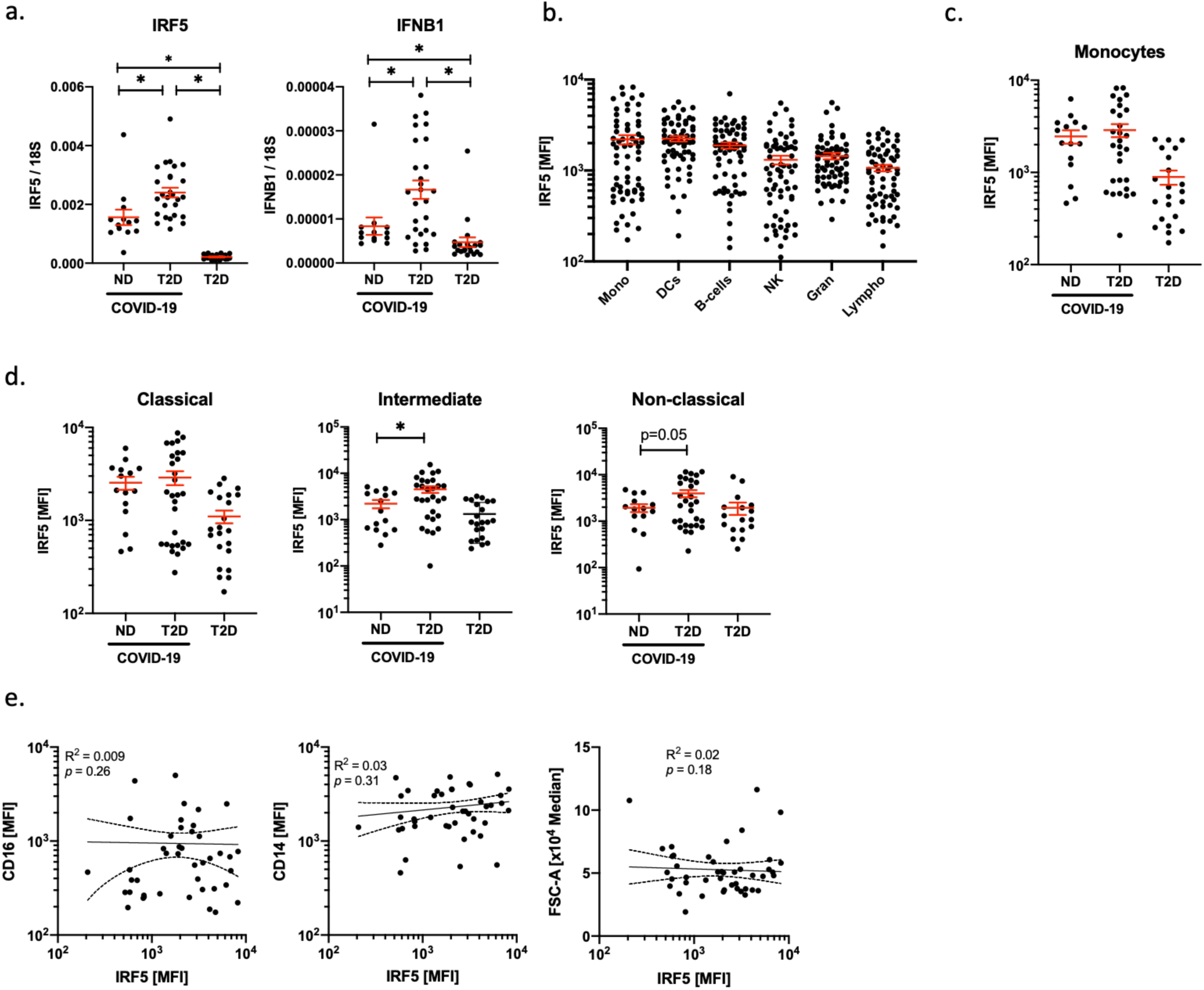
Characterisation of monocyte morphology and inflammatory cells from peripheral circulation ND and T2D COVID-19 patients. Related to figure 3. IRF5 and IFNB1 mRNA expression in COVID-19 patients with type-2 diabetes (T2D) and in non-diabetic (ND) patients (a). IRF5 expression by flow cytometry in different immune cell populations found in circulation in ND and T2D COVID-19 patients (b). IRF5 expression by flow cytometry in monocytes of ND and T2D COVID-19 patients and in T2D patients without COVID-19 (c). IRF5 expression by flow cytometry in monocyte subpopulations of ND and T2D COVID-19 patients and in T2D patients without COVID-19 (d). Correlative analyses of IRF5 expression by flow cytometry, to the expression of CD14, CD16 and to monocyte size (FSC) in monocytes of ND and T2D COVID-19 patients (e). Differences between groups were evaluated with unpaired t-test. Analyses of variance (ANOVA) or covariance (ANCOVA) were used for multiple group comparisons. For correlative analysis Spearman’s test was carried out calculating a 2-tailed *p*-value. * : p < 0.05

**Figure S4.**
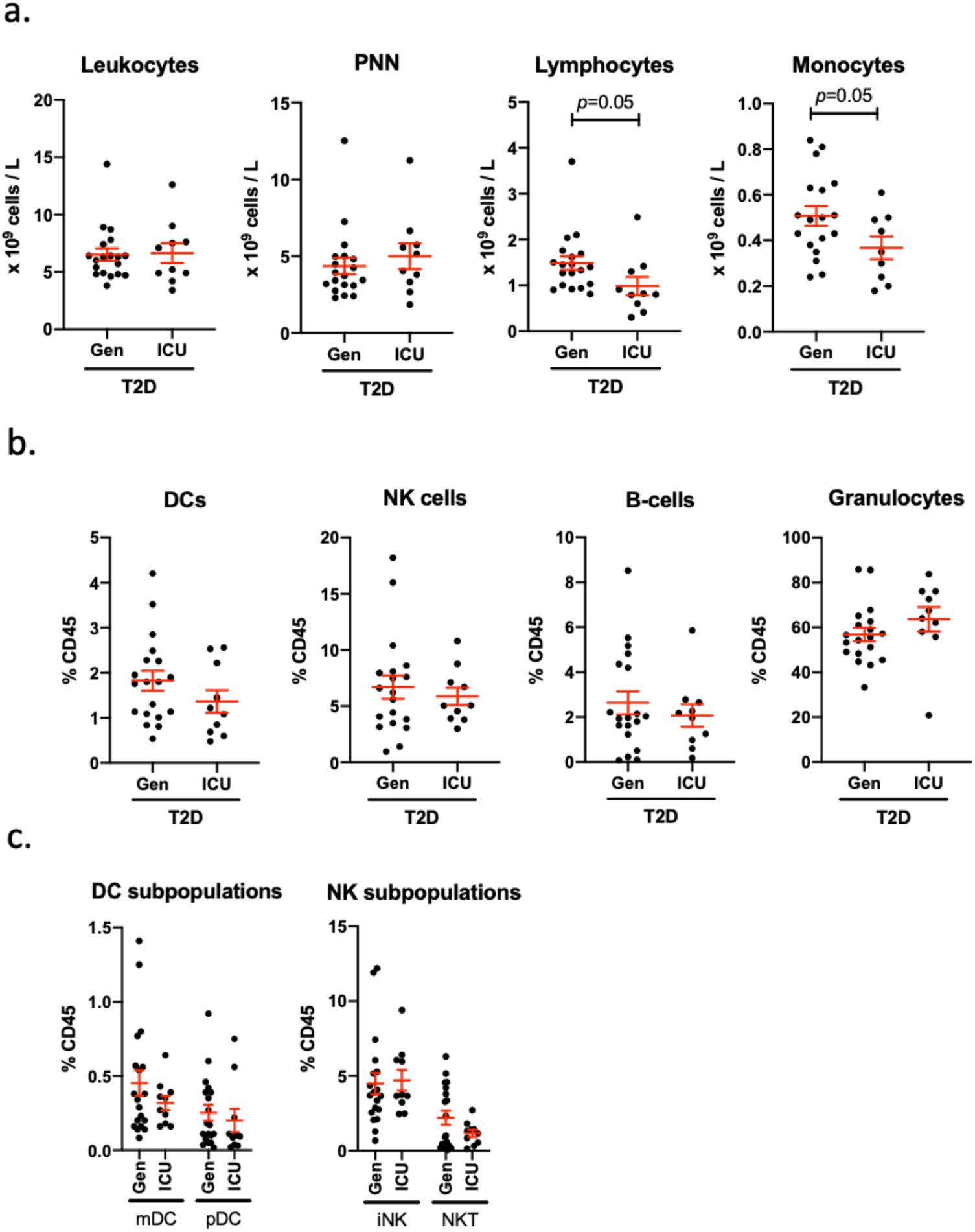
Immunophenotypic analyses of T2D patients with COVID-19 treated in general wards or in ICU. Related to figure 4. Absolute quantification of circulating leukocytes from full blood counts at admission of type-2 diabetic (T2D) COVID-19 patients treated in general wards (Gen) or in the intensive care unit (ICU) (a). Flow cytometric quantification of dendritic cell (DC) natural killer (NK) cell, B cell and granulocyte frequency in peripheral blood of T2D COVID-19 patients treated in Gen or in the ICU (b). DC and NK subpopulation frequency in peripheral blood of T2D COVID-19 patients treated in Gen or in the ICU (c). Data are presented as mean +/− SEM. Differences between groups were evaluated with unpaired t-test.

